# Automated Annotation of Disease Subtypes

**DOI:** 10.1101/2023.09.24.23296020

**Authors:** Dan Ofer, Michal Linial

**Affiliations:** Department of Biological Chemistry, The Life Science Institute, The Hebrew University of Jerusalem, Israel

**Keywords:** Disease subtypes, Disease ontology, Explainability, Machine learning, Medical language models, Ontology completion, Open Targets, Orphanet, Personalized medicine

## Abstract

**Background:** Distinguishing diseases into distinct subtypes is crucial for study and effective treatment strategies. The Open Targets Platform (OT) integrates biomedical, genetic, and biochemical datasets to empower disease ontologies, classifications, and potential gene targets. Nevertheless, many disease annotations are incomplete, requiring laborious expert medical input. This challenge is especially pronounced for rare and orphan diseases, where resources are scarce.

**Methods:** We present a machine learning approach to identifying diseases with potential subtypes, using the approximately 23,000 diseases documented in OT. We derive novel features for predicting diseases with subtypes using direct evidence. Machine learning models were applied to analyze feature importance and evaluate predictive performance for discovering both known and novel disease subtypes.

**Results:** Our model achieves a high (89.4%) ROC AUC (Area Under the Receiver Operating Characteristic Curve) in identifying known disease subtypes. We integrated pre-trained deep-learning language models and showed their benefits. Moreover, we identify 515 disease candidates predicted to possess previously unannotated subtypes.

**Conclusions:** Our models can partition diseases into distinct subtypes. This methodology enables a robust, scalable approach for improving knowledge-based annotations and a comprehensive assessment of disease ontology tiers. Our candidates are attractive targets for further study and personalized medicine, potentially aiding in the unveiling of new therapeutic indications for sought-after targets.

## 1. Introduction

Disease subtyping, also called disease stratification, enables a more precise understanding and characterization of various illnesses, paving the way for personalized treatments and improved patient outcomes. Disease subtypes can be delineated using genetic, molecular, or clinical attributes [1]–[3].

As personalized medicine advances, disease subtyping can advance our understanding of disease mechanisms across various medical disciplines [3], [14]. Moreover, it is needed for study, effective treatment, and discovering potential cures. Furthermore, certain drugs and treatments may be relevant only for specific subpopulations and disease manifestations [15], [16]. Disease progression can also be markedly different, requiring different clinical treatment regimes [17]–[19].

We concentrate on clinically significant differentiation, or subtyping, of diseases. For instance, variants of SARS-Cov-2 caused the COVID-19 pandemic. We claim that subvariants (e.g., delta, omicron) are not useful for clinical categorization. Instead, the partition of COVID-19 to patients experiencing an acute phase and others who exhibit persistent conditions known as long COVID dictates clinical importance. Another example of disease subtyping is evident in the differentiation between type 1 and type 2 diabetes mellitus, where, despite the similarity in dysregulation of blood sugar levels, treatment approaches, disease management, and potential cures vary significantly. Neurodegenerative disorders like Alzheimer’s and Parkinson’s, although categorized clinically as neurodegenerative diseases, exhibit distinct molecular pathologies, subtypes, and diverse progressions and treatments. Advancing our understanding of Parkinson’s subtypes is pivotal for devising effective treatment strategies [1].

Conversely, while various viruses can cause influenza, differentiating them based on their specific causal virus is clinically irrelevant since treatment and disease progression remain identical.

Historically, the medical community has spearheaded efforts to identify the multifaceted nature of diseases. Clinicians primarily rely on the International Classification of Diseases (ICD), which undergoes periodic revisions [4]. For instance, Diabetes mellitus (ICD-10, E10-E14) is partitioned into Type 1 (E10), Type 2 (E11), and unspecified diabetes (E14), along with further subtypes [5], [6].

The Open Targets (OT) platform integrates a variety of molecular, genetic, and biomedical datasets, ontologies, and knowledge graphs [7]. Increasing quantities of semantic resources offer a wealth of knowledge but also increase the probability of wrong knowledge-based entries and error propagation [8]–[11]. Thus, developing automated approaches to both complete and correct potentially spurious entities in large knowledge bases is of paramount importance. The concept of accurate hierarchical categorization of diseases and phenotypes is further underscored by initiatives like the gene ontology (GO) project, where ontologies, phenotypes, and functions across species are mapped to coding genes [12]. The impact of the Gene Ontology (GO) project on automatic functional annotation tasks such as CAFA is unquestionable [10], [13]. CAFA (Critical Assessment of Functional Annotation) is an ongoing effort to evaluate and improve the computational annotation of protein functions.

Existing ontologies are complex and may suffer bias due to many factors, including population prevalence and the number of researchers and clinicians working on the disease, factors that may impact their division into sub-categories in the literature as well as the quality of annotation [20]–[22].

Most existing methodologies rely on inheriting or directly mapping disease levels from existing annotations and ontologies by strict, manually defined rule-based methods. One concentrated on a narrower domain, clustering specific cancer data, imaging, and non-biomedical data [23]. It did not endeavor to offer predictions across a broad spectrum of known diseases. Another approach used by OT data is to identify drug-disease associations, which is a different objective from ours [24]. Our work also relates to knowledge-based link prediction, and literature-based discovery [25]–[29]. However, these mainly aim to identify “horizontal links” between existing topics. In contrast, our objective is to flag topics that might have undiscovered subtopics, or missing “vertical” links.

We propose a data-driven machine learning approach for ontology completion and correction, specifically applied to OT. OT integrates a wide variety of gold-standard curated ontologies and data sources, from which we curate a novel benchmark dataset for disease subtype prediction. This dataset can be used for evaluating and developing approaches for characterizing diseases. Furthermore, we present an approach for identifying and evaluating candidate diseases with potential novel subtypes and mis-annotations, and a ranked list of predictions. We validate our novel candidates using ongoing research and future OT annotation updates. Our automated approach is interpretable, scalable and offers novel candidate disease subtypes for future research.

We outline the key steps in our approach. First, we create a target matrix from the existing OT ontology for all diseases, defined as whether a disease has a subtype or not. Predictive features for each disease are derived from OT’s direct evidence data sources. A machine learning model is trained on known targets. Predictions are formulated for every entry in the dataset through iterative rounds of hold-out cross-validation. Subsequently, we interpret and scrutinize the results and models. Instances where the predicted target consistently deviates from the known one, coupled with supplementary filtering, are identified as potential candidates for novel subtypes or highlight annotation inaccuracies. Our goal is to help find unknown disease subtype candidates within existing databases.

## 2. Results

### 2.1 Diverse Disease Ontologies

A machine learning model for disease subtyping assessment and discovery was developed using the sources integrated into OT. An overview of these sources is demonstrated in **Fig 1**.

**Fig 1.**
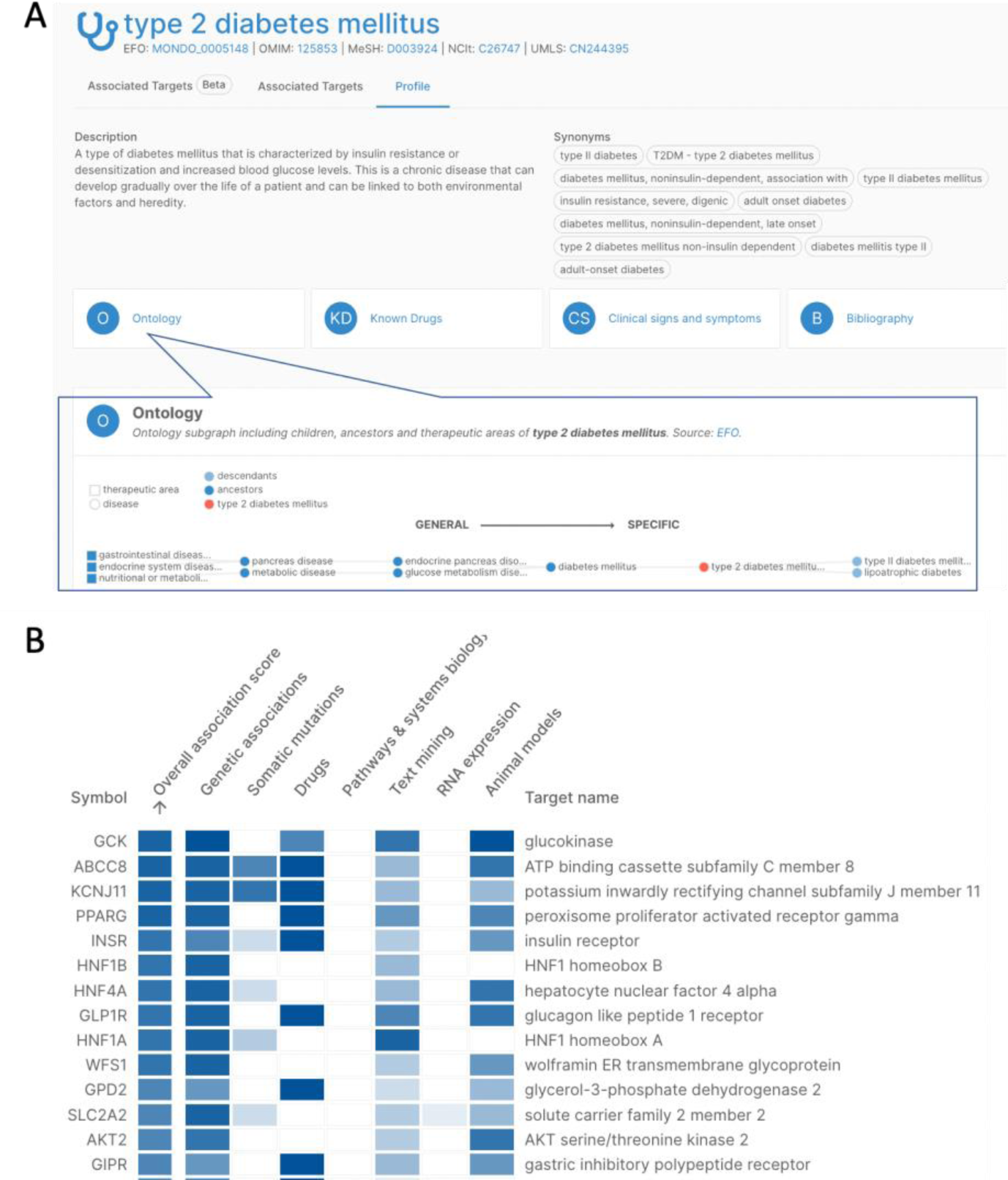
Example from Open Targets Platform for type 2 mellitus diabetes. **(A)** Description, disease ontology (including any subtypes), synonymous nomenclature, known drugs, bibliography, and clinical symptoms **(B)** Weighted evidence sources and domains for the disease, including genetics (genetic associations), somatic mutation, drugs, text mining, and more. Each column is colored by the intensity of the relevant score (normalized 0 to 1). The gene list is sorted by the overall association score.

**Fig 1A** is a disease perspective view of type 2 diabetes mellitus on the OT platform [30]. It encompasses text associated with the description, synonyms from various databases, and summary statistics for additional information, including ontologies, known drugs, clinical signs, symptoms, and bibliography. **Fig 1B** lists associated genes for type 2 diabetes mellitus ranked by global scores for the disease. The genes’ evidence is indicated by the heatmap, with genetic association from genome-wide association studies (GWAS), direct support from drugs, text mining, RNA expression, animal model studies, and more.

The final dataset held 17,222 diseases, of which 5,848 (34%) have known subtypes. Feature importance and model performance were evaluated in predicting targets with known subtypes. Our novel features demonstrate high importance and predictive power. In addition, these may support the discovery of novel therapeutic indications for highly pursued targets.

### 2.2. Performance Evaluation - Known Targets

For the task of predicting known disease subtypes, we tested multiple machine-learning models, including logistic regression (LR), random forest (RF), CatBoost (a boosting tree model), as well as domain-specific baselines. Binary classification performance was evaluated using five-fold stratified cross-validation [31]–[33] (**Fig 2)**. Additional evaluation results are provided in Supplementary table S1, and the confusion matrix of the best, CatBoost, model in Supplementary Fig S1.

**Fig 2.**
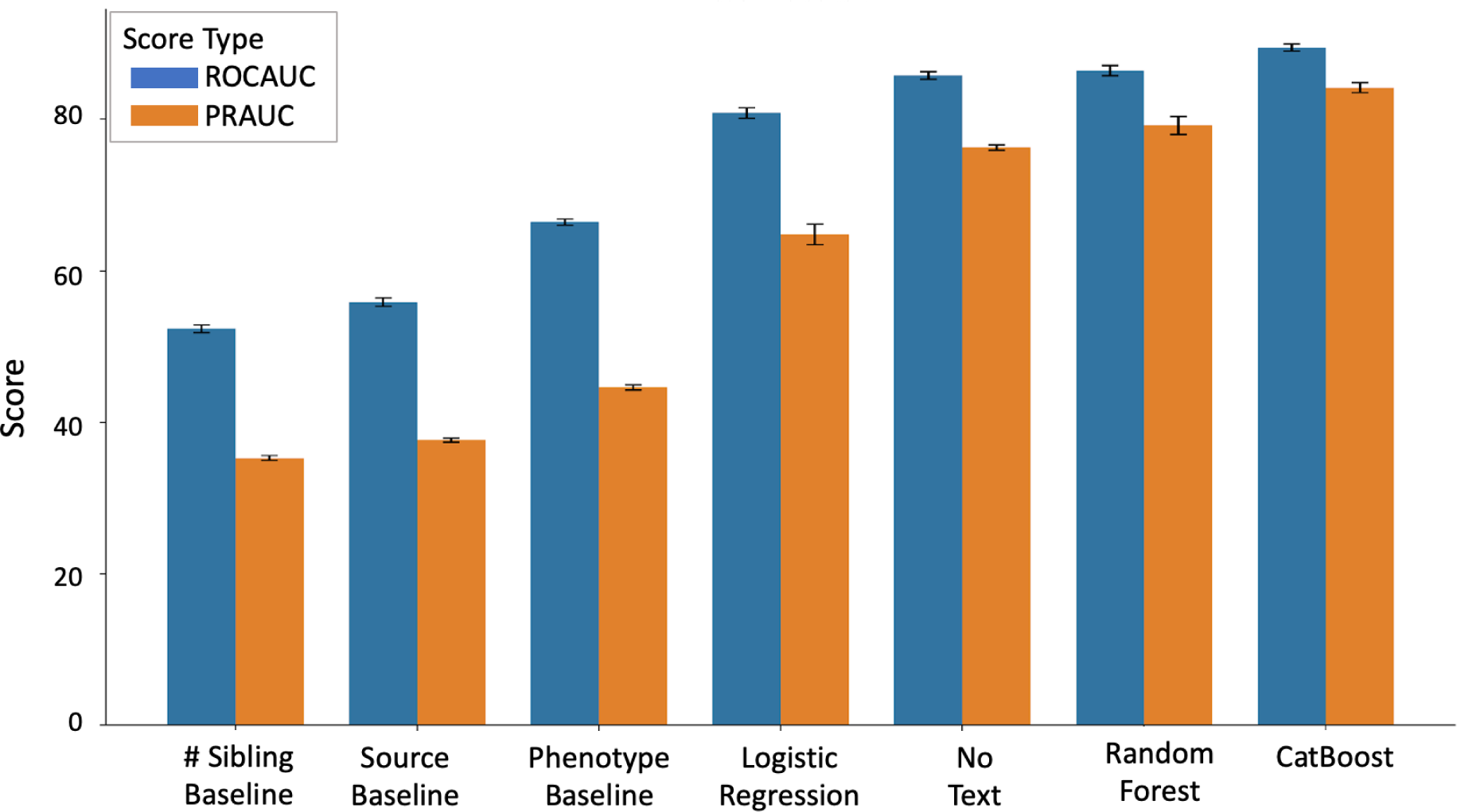
Known subtypes model evaluation. Results shown 5-fold cross-validation, and standard deviation, for a selection of evaluated models. PRAUC: Area Under Precision-Recall Curve. No Text - CatBoost model with text embedding features excluded (see Methods: “Deep learning using Text features”). The remaining, non-baseline models used all features. Additional models and metrics results are reported in Supplementary table S1.

For comparative analysis, we added to the assessment three domain-specific baselines. These are linear models trained exclusively on a single feature: (i) the disease’s database source (e.g., Orphanet); (ii) the number of known phenotypes (“phenotype frequency”); (iii) The number of “siblings” a disease has in OT database, wherein all “siblings” share the same parent disease. All baselines outperform random guessing. All models substantially outperform the baselines (**Fig 2**). CatBoost had the best performance, achieving a ROCAUC (Receiver Operating Characteristic Area Under the Curve) of 89.4% (**Figs 2-3**). Accordingly, we used CatBoost for subsequent predictions of novel subtypes and analyses. This included extracting novel predictions and ablation analysis of the text features. We find that text features significantly enhanced performance when compared to a model devoid of text features (**Fig 2**, “No text model”), yielding an AUC of 0.89, in contrast to 0.86 without these features.

**Fig 3.**
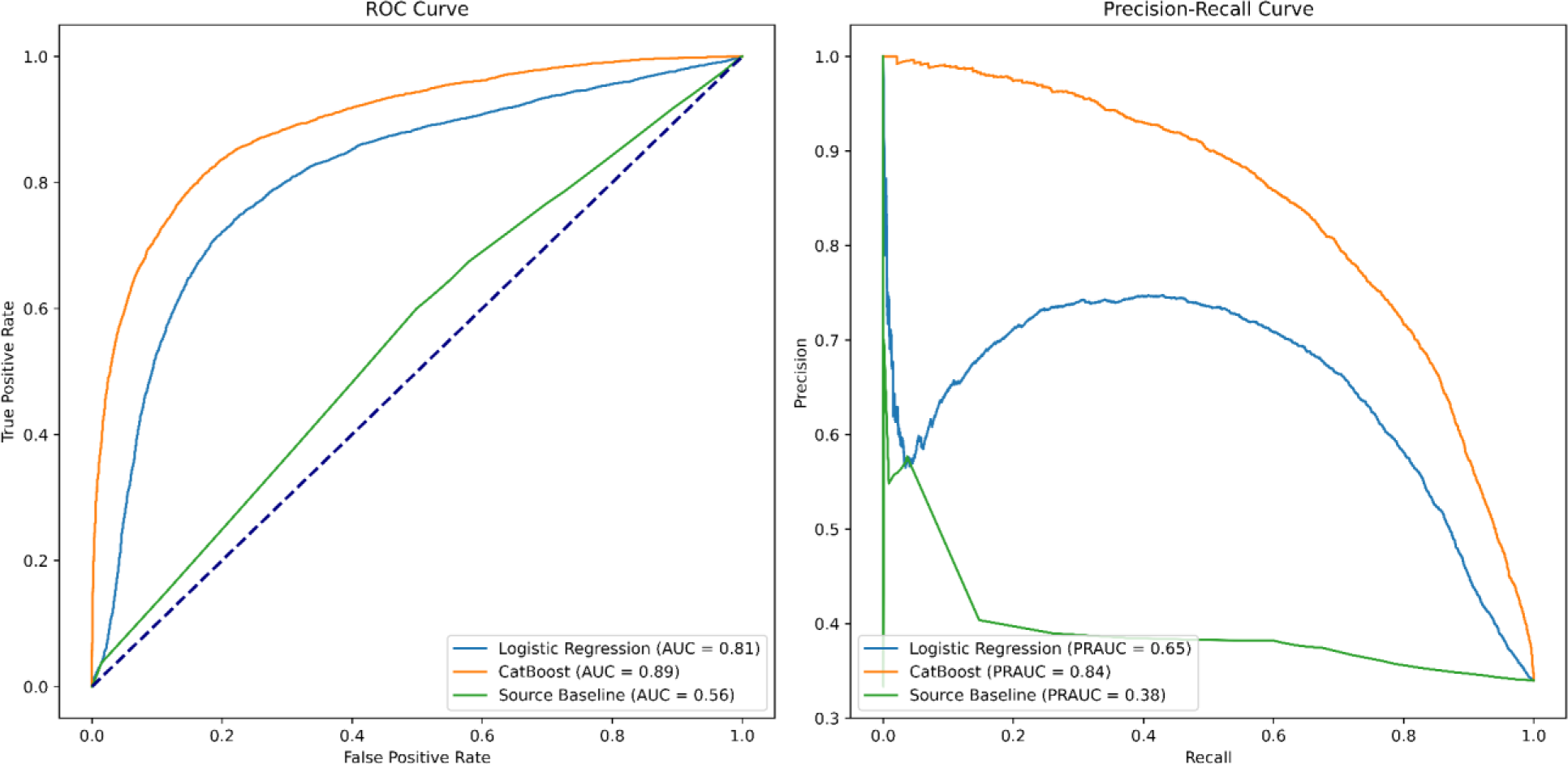
Known subtype models comparison. Model test-set predictive performance is measured by ROCAUC (left) and PRAUC (right).

### 2.3. Model Feature Importance

The “known subtypes” model’s feature importance was extracted using SHAP **(Fig 4A)**. We observe that the source database is a major feature, as we might expect (e.g., Orphannet diseases are more likely to be understudied and to lack subtypes). Diseases with a high amount of genetic and literature evidence score (**Fig 1**) are more likely to have subtypes. Specific disease phenotypes were strong features in aggregate, but consisted of hundreds of individual weaker features, and thus are not visible here; the engineered feature of the highest global frequency of an associated phenotype (“Max phenotype frequency”) is strong - we theorize it might help the model learn about diseases with easy versus hard to characterize phenotypes. The number of phenotypes is another interesting feature. disease with many different effects may be more complicated to stratify or maybe a combination of effects. The various text features from the pretrained biomedical large language model have a clear impact (see Methods: “Deep Learning Text Features”). These might help extract additional information about diseases from their descriptions or pre-existing literature.

**Fig 4.**
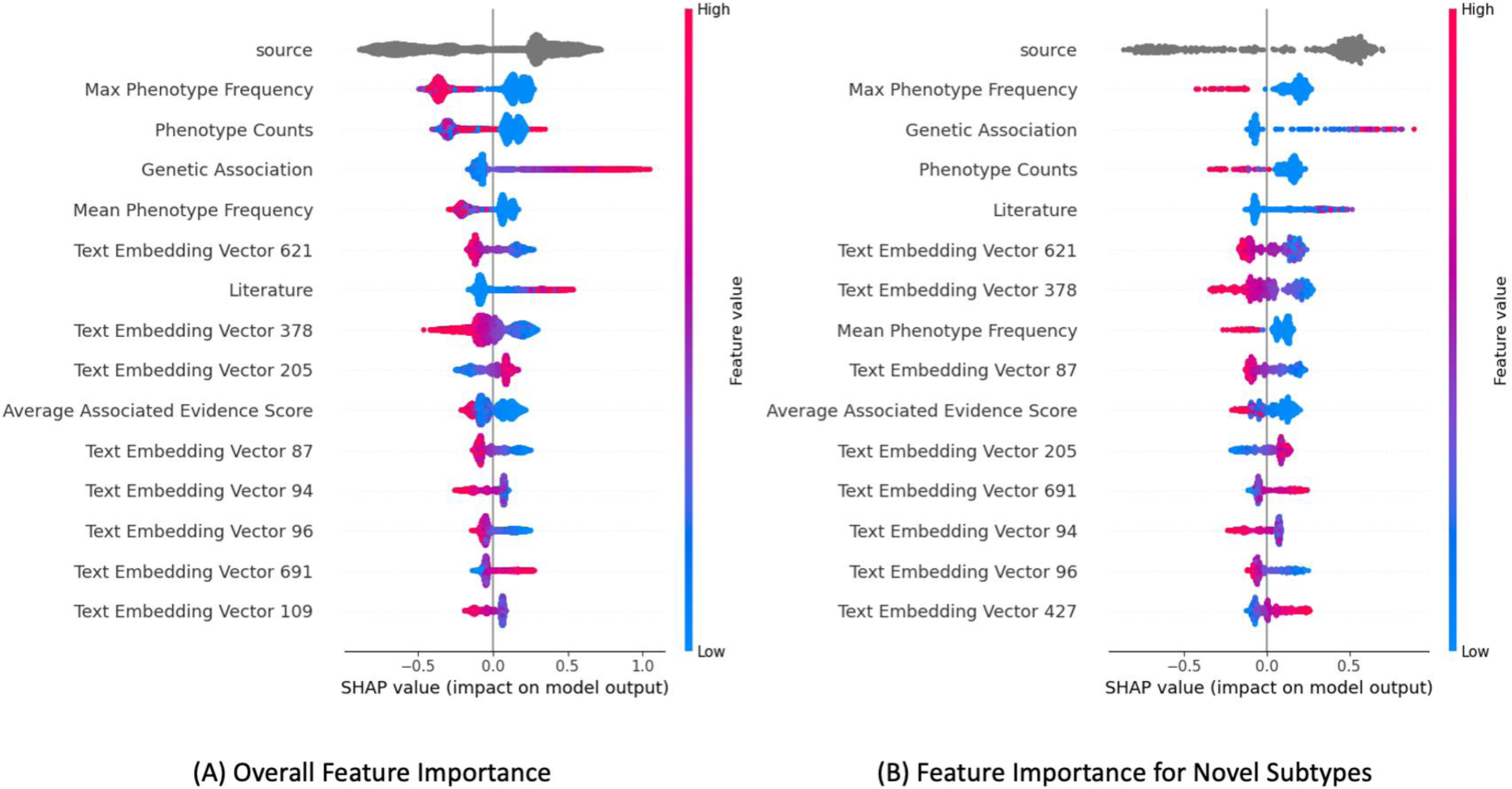
Feature importance over data subsets. Shapley-based feature importance in disease subtype prediction for A) All data (Existing OT subtype annotations). B) Feature importance for subset with predicted novel subtypes, and no subtype in ground-truth. Top 15 features shown. See Methods for feature dictionary.

### 2.4. Predicting Diseases with Novel Subtypes

Given the strong performance of machine learning models in our evaluation, we used our model to predict disease subtypes for diseases that are not identified as such in the existing dataset. We applied a repeated-stability approach to identifying potential novel candidates. We report on cases where a model, retrained over multiple random data splits (using 8×5 repeated stratified cross-validation), consistently (eight out of eight times) predicts a different label than the recorded one per data point. The predicted data point is always part of the held-out test set. Thus, there are 8 held-out model predictions for every instance in the data. We identified 1,546 such cases, out of 17,222 records in the dataset. Of these, 515 (33%) are predicted to have subtypes, where none are recorded in the OT ground truth. The average prediction consistency was 84.9%. This approach is effectively an ensemble. In supplementary S5 we record the averaged model predictions on the held-out test set splits. This ensemble has better results than a single model, as might be expected (ROCAUC: 91.08, PRAUC: 86.61) [19], [34]. Thus, we used these as candidate predictions. The full list of candidate predictions is in Supplementary File S5.

Using SHAP we examined features’ contributions to model predictions, for the predicted novel subtypes subset only **(Fig 4B)**, and observed that the top features (evidence source, phenotype etc) remain relatively stable in terms of rank importance and direction of effect, with the same effect as for known subtypes in the general population (**Fig 4B**), indicating that these cases are not anomalous in terms of their features compared to the background. We plotted the distribution of several “top” (selected by model importance) features, and observed a similar distribution overall (**Fig 5)**, again reinforcing that the novel subtypes have similar properties to the ground-truth known subtypes.

**Fig 5.**
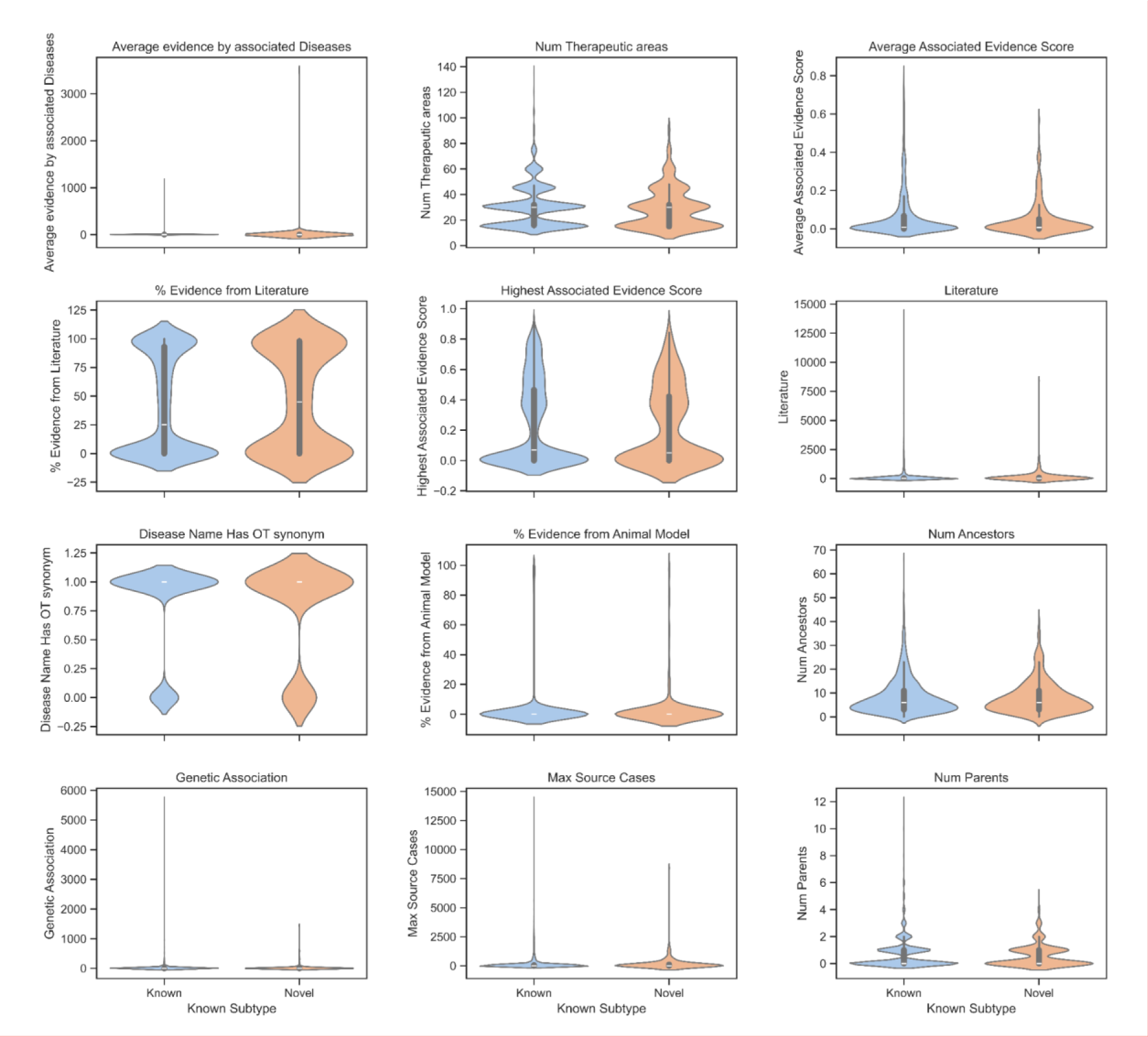
Distribution of known vs novel subtypes. Violin-plots of selected features for 5.8K diseases with **known** subtypes vs. 531 predicted, candidate **novel** subtypes that have no known subtype.

### 2.5. Evaluation of Predictions for Unknown Targets

As there is no ground truth to evaluate the novel subtype predictions, we use the scientific literature as an external validation mechanism, as in [24]. We presume that potential subtypes might be discussed in the wider literature before any validation and integration into existing knowledge. From our models, we selected 800 predictions, sorted by highest predicted probabilities, evenly split with 400 cases per target class. We searched for literature hits of these candidate diseases being mentioned as having a subtype, sub-manifestation, or pleiotropy in PubMed. We extracted the fraction of such cases relative to the total number of literature references for each disease (Supplementary Table S3). The difference between candidates categorized by their predicted subtype was statistically significant as determined by a one-sided unequal variance t-test (*p-value = 2.6e-7*), with predicted candidates having ∼5 times as many results with subtypes (i.e., 2,100 vs. 400).

We conducted a similar analysis on 300 of the stable predicted novel candidates (Supplementary Table S4), specifically those whose predictions deviated from existing annotations. For this subset, the literature search revealed no significant difference (p-value=0.27). This aligns with our hypothesis that these novel candidates are uncharacterized in existing studies. If our model’s predictions were merely identifying diseases widely acknowledged to have a subtype but not yet annotated in OT, this would be evident by the literature search (i.e., numerous “subtype” mentions), which was not observed here. This observation bolsters our assertion that the model is highlighting truly unknown subtype candidates, rather than just inadequate or faulty annotations.

#### 2.5.1 Temporal Validation of Candidates

For further validation that our candidate predictions are meaningful, we downloaded an additional, 18-month newer snapshot of the OT database (dated 12.2023). Of the 19,819 diseases that overlap in both versions (the total population), 1.2% (238) had subtypes added or removed entirely between versions. 1509 (92%) candidates were successfully matched by name, and of these, 48 (3.18% of the candidate sub-population) had their subtype annotations changed. The 3-fold proportional difference (3.18% vs 1.04% in the sub-population not containing candidates) is statistically significant (p-value <1e-05, one-way, two proportion z-test). This shows that our candidates are far more likely to have their existing annotation ground truth “fixed” in accordance with our predictions, relative to the overall population.

#### 2.5.2 Analysis of Database Source Distribution in Predictions

A concern was whether the model was simply identifying surface-level patterns, such as associating all diseases from a specific data source, such as Orphanet (an orphan disease database) with a subtype. Such hidden confounders are common in many predictive scenarios [35], [36]. To validate the model, we examine the distribution of disease subtypes in our predictions against the known subtypes, focusing on their source database (**Table 1**). We find the distribution of our novel candidates slightly differs from the original dataset target at the database level. Notably, there is a lower frequency of subtypes in predictions from Orphanet. This gives further support to our identification of candidates using non-trivial patterns, and reducing bias towards existing annotation sources.

**Table 1.**
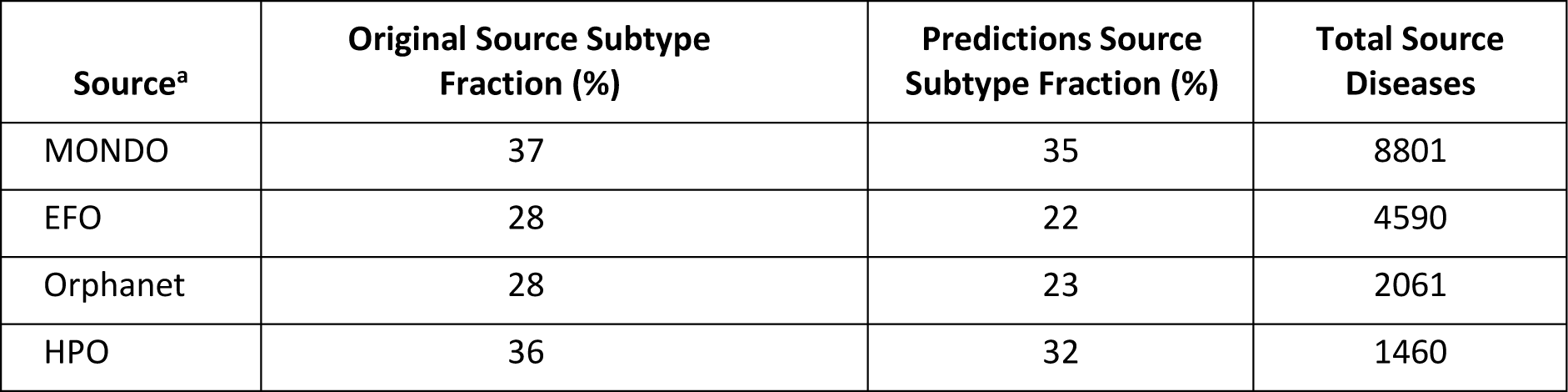
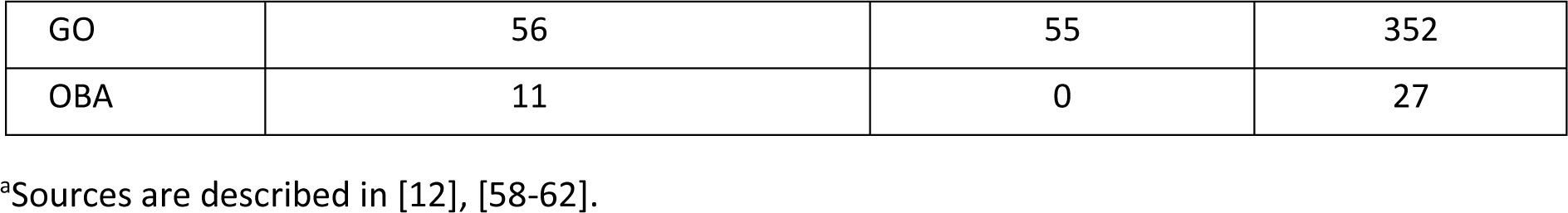
Source subtype distribution.

**Table 1** shows the distribution of diseases with known vs. predicted subtypes, grouped by the 6 largest database sources. Diseases’ subtype fraction is shown per source. “Original Source Subtype Fraction” depicts the percentage of diseases with a subtype in existing annotations. “Predictions Source Subtype Fraction” depicts the percentage of diseases with predicted subtypes in novel predictions. “Total Source Diseases’’, indicates the total number of diseases (regardless of subtype) from a source.

#### 2.5.3 Understanding Potential Novel Disease Subtypes

Following our model’s identification of 515 diseases predicted to have subtypes not currently annotated in OT, understanding the significance of these findings becomes paramount. These candidates, selected based on their consistent predictions and absence of known subtyping, underscore the vast potential for refining our understanding of disease taxonomy. The top-ranked novel predictions were manually reviewed. We provide several explanations for representative novel candidates **(Table 2)**. Broadly, we note high-level causes that include: (P) Pleiotropic manifestations: different causes resulting in seemingly similar outcomes, leading to diseases with varied presentations. Additionally, overlapping clinical presentations can cause misdiagnosis. Examples include the confusion between CNS inflammatory disorders and multiple sclerosis (MS), neurodegenerative diseases and dementia [17–18,40]. (B) Variability in disease course and treatment: Clinical trajectory and therapeutic responsiveness can vary based on disease subtypes and interaction with patient characteristics (e.g., Parkinsons’ [1]). (H) “High-level” semantic terms: these are inherently broad and include a range of conditions, e.g., “infections”. Such cases are clear-cut and may be due to a lack of linkage of known terms between ontologies.

**Table 2.** Candidate novel subtypes’ explanation by categories.

| Disease | Explanation | <sup>a</sup>Rationale | Ref |
| --- | --- | --- | --- |
| COVID-19 | The spectrum of clinical presentations, from asymptomatic states to severe respiratory syndrome, and long-term impairment ("long COVID") suggests potential subtypes. | B | [42],<br>[43] |
| Smallpox | There are four types: ordinary; modified (mild, occurring in previously vaccinated persons); flat; and hemorrhagic. For example, the Smallpox vaccine does not protect against hemorrhagic smallpox. | B | [63] |
| Female breast carcinoma | Varied molecular subtypes, defined by specific gene expression profiles, as well as anatomical regions, dictate prognosis and therapeutic responsiveness. | P, B |  |
| Nephropathy | Kidney pathologies with distinct histological features and clinical courses. | P, B |  |
| Cardiovascular risk | Disease families with broad risk factors, including environmental, genetic and genetic-environment interaction (GxE). For example, diabetes, or specific genetic risk factors. | P, B | [6],<br>[15] |
| Structural epilepsy | Different brain anomalies can precipitate varied forms of epileptic seizures. It may coexist with tumors, cysts, stroke, or vascular malformations. | P, B | [41] |
| Parkinson disease, mitochondrial | Neurodegenerative disorders, that though clinically overlapping, have distinct molecular pathologies, and treatment.. | B | [1] |
| (Multiple) malignant and non-malignant tumors | Behavior, complications and treatment are influenced by specific mutations, patient genetics and risk factors (GxE), physical size, organ location. | H |  |
| Eye infections | Distinct etiological agents, spanning bacteria, viruses, fungi, and parasites, involving distinct treatment and risks. | H |  |
| Alcohol dependence;<br>Sexual and gender identity disorders;<br>Speech disorder;<br>Central nervous system development | Semantically high-level categories, lacks breakdown to a clinically useful level, required for effective treatment | H |  |
| Dysplasia | A high-level family of conditions (encompassing “types of abnormal growth or development of cells, organs”, and resulting abnormalities. Different categories by delineation by microscopic (cell) level, organ (macroscopic), organ and cell type. | H | [44] |

<sup>a</sup>Rationale: High level rationale codes: (P) Pleiotropic manifestations. (B) Variability in disease course and treatment. (H) “High level” semantic terms.

In the cases of diseases predicted to be misannotated and to not have a subtype; some cases may simply be model errors, hence the need for a final layer of expert review. There are numerous valid explanations for why a disease may have an incorrect annotation, ranging from human error, database error, and annotator guidelines biases. We illustrate it by a hypothetical case of two different virus strain variants of the SARS-CoV-2 Omicron strain being classified as two distinct diseases. While they are caused by a separate strain, their disease manifestation and course of treatment overlaps with that of a flu-like illness. In this case, we claim that the distinction is not clinically meaningful, but could be recorded as such by mistake. Merging such subtypes would improve the database. Our temporal validation showed that our candidates are much more likely to be “interesting” and in need of reassessment in the OT database.

In the cases of diseases predicted to be misannotated and to not have a subtype; some cases may simply be model errors, hence the need for a final layer of expert review. There are numerous valid explanations for why a disease may have an incorrect annotation, ranging from human error, database error, and annotator guidelines biases. For example, a hypothetical case of 2 different variants of Omicron strain of SARS-CoV-2 being classified as two distinct diseases. While they are caused by a separate strain, it is the common state of sessional flu. In this case, we claim that the distinction is not clinically meaningful, but could be recorded as such by mistake. Merging such subtypes would improve the database. Our temporal validation showed that our candidates are much more likely to be “interesting” and in need of reassessment in the OT database.

#### 2.5.4 Understanding an Individual Prediction

We present an illustratory model explanation example for a novel predicted subtype candidate, COVID-19 (**Fig 6**), using the known subtypes model’s SHAP explanation.

**Fig 6.**
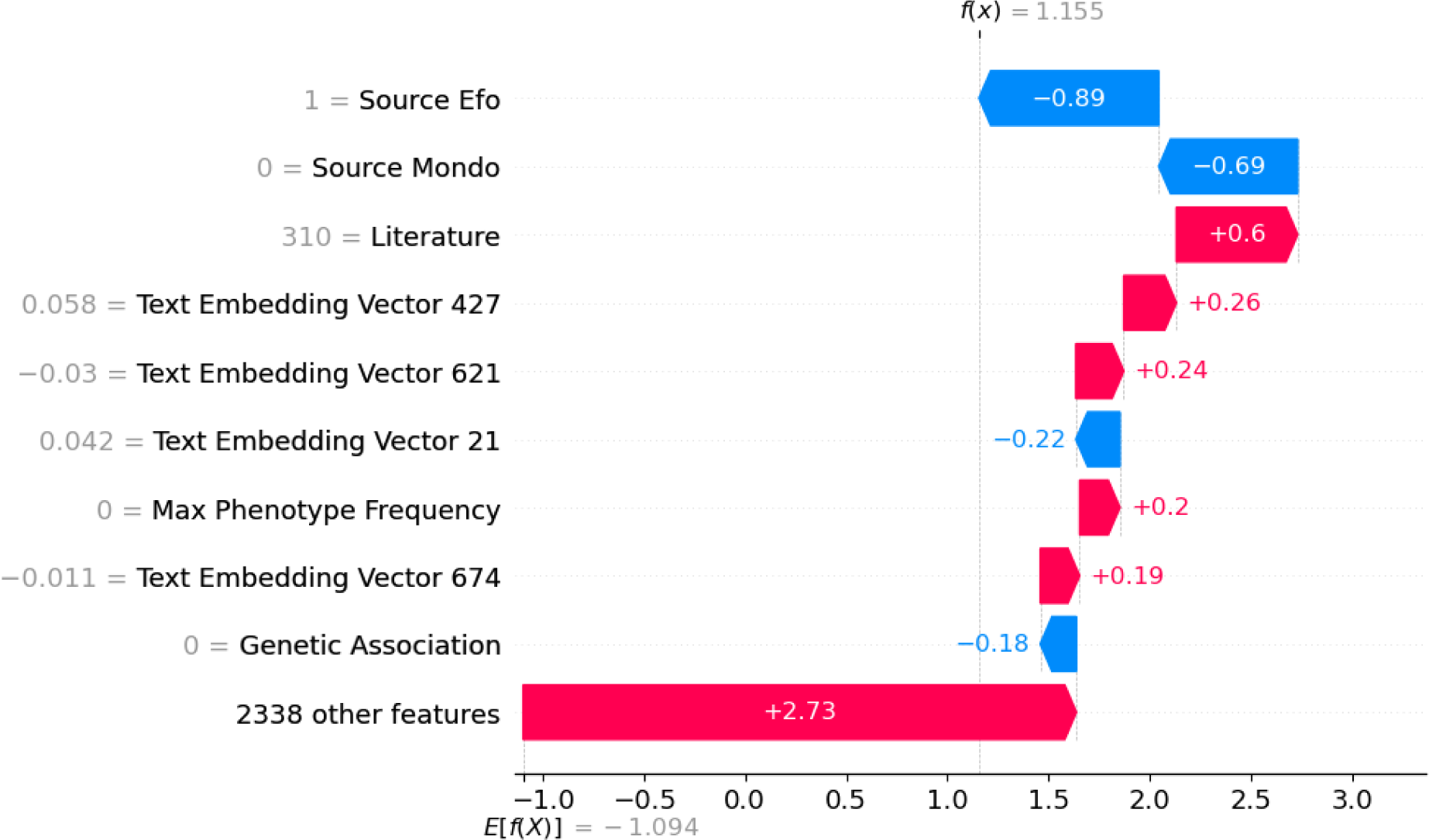
SHAP explanations for COVID-19. Explanation of a single positive (“1” - has subtype) prediction as a SHAP waterfall plot. The SHAP value of a feature represents the impact of the evidence provided by that feature on the model’s output. The waterfall plot shows how the SHAP values (evidence) of each feature move the model output from the prior expectation under the background data distribution, to the final model prediction given the evidence of all the features. Colour and direction of the arrows indicates the direction of effect. Feature values shown on the left in grey.

Examples of exemplar disease text excerpts with of high, neutral and low values per each of the embedding dimensions (see 4.4) are shown (**Table 3**)

**Table 3.**
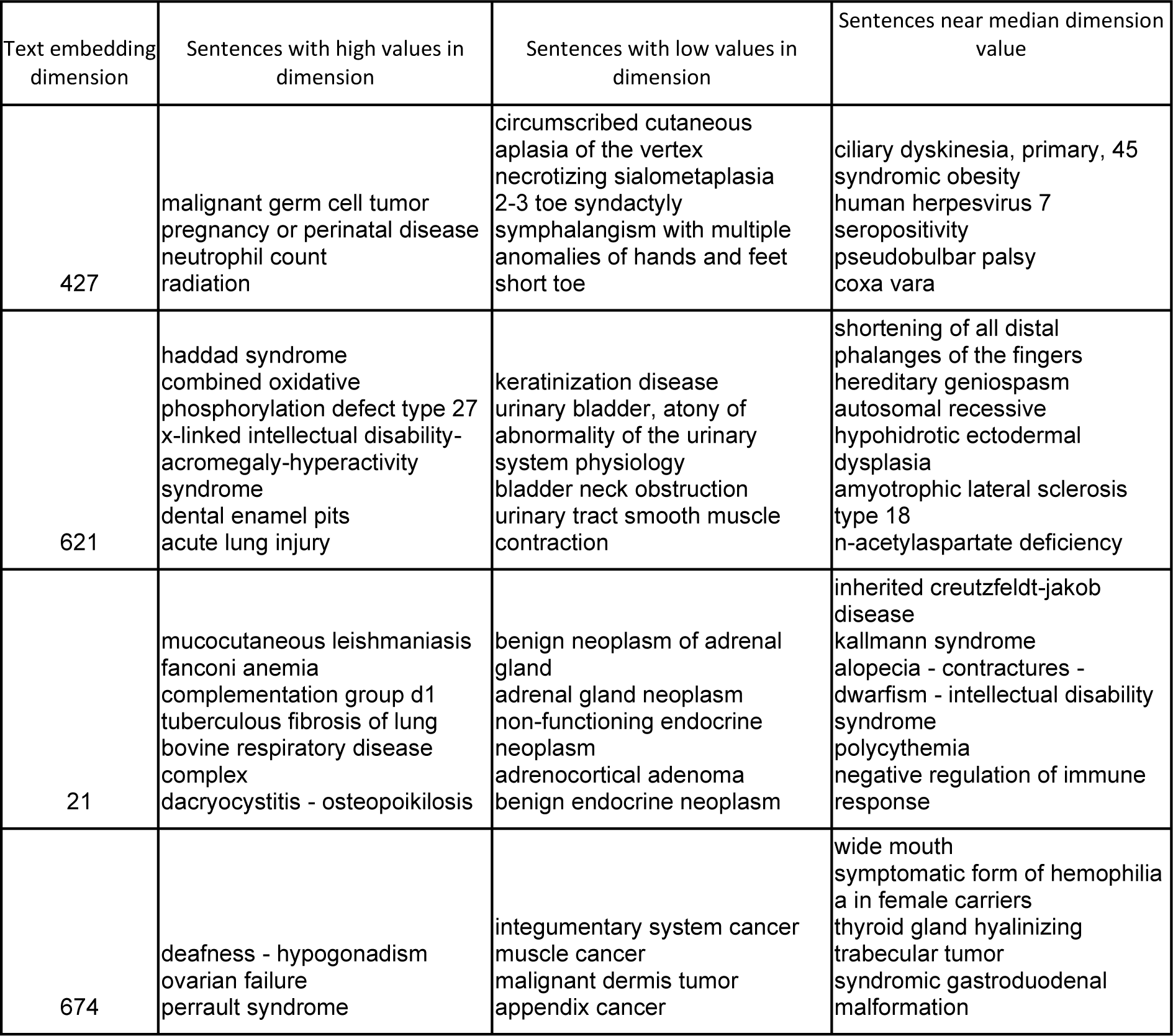
Text embedding explanations for COVID-19.

## 3. Discussion

Discovering disease subtypes is an important problem in medicine, with applications in both basic research and personalized treatment. That a disease may even potentially have subtypes is not an obvious fact. Historically, diagnoses like “female hysteria” led to ineffective treatments, overshadowing the recognition of genuine diseases or conditions [37]. It wasn’t until later that such broad diagnoses were deconstructed into specific diseases. Parkinson’s Disease, which can be caused by drug toxicity or vascular malfunction, is another major example. There is considerable work into finding subtypes that can help treat patients and predict the future course of the disease’s progression [1], [17].

Our hypothesis posits that diseases with distinct subtypes are discernible based on intrinsic aspects of the disease itself as well as meta-features relating to its research. Consider cancer diseases that are mostly driven by somatic mutations as opposed to predisposition germline genetic variants. Single-gene disorders are often inherited but they may split into early and late onset diseases that might be addressed differently in clinical terms. In this aspect, early and late onset of diseases are documented to Alzheimers [67]), Parkinson [1],[68] and numerous autoimmune conditions (e.g., Crohn’s disease, myasthenia gravis). Thus, we focused our features on representing these aspects of different diseases. We also address “meta-science” aspects about diseases, such as their research process, and limits on studying them. For example, diseases with many distinct animal models (as reported by OT), extensive literature and many candidate drugs, are less likely to be categorized as “orphans” than those observables only in humans. Furthermore, some phenotypes are easier to observe, measure and categorize, while others may be more nuanced. Obvious cases include developmental disease causing facial deformities vs mental health conditions. Our features help the models learn these various aspects, in a way which aims to be largely objective, with the goal of learning from the intrinsic characteristics of the diseases themselves, as opposed to approaches that might be more biased towards existing literature and annotations, such as text mining [34], [38], [39]. This approach can yield better predictive performance than underlying, partial annotations or rule-based systems, as has been observed in other works, such as healthcare mortality prediction[19].

Possible confounders are the rarity of diseases. This can be partially quantified by the disease’s population prevalence, using the UK biobank (UKB) [40] population frequency. Another metric of rarity is the classification of a disease as an “orphan disease”, which is determined by its source (e.g., listed in Orphanet). Interestingly, the UKB calculated prevalence has low feature importance, and is not an impactful feature to the model predictions (Fig 4). Its removal did not affect model evaluation results (not shown), further emphasizing that our task is indifferent to it. Thus, we disqualify it as a confounding proxy for model performance. On the other hand, the source feature is important overall. We evaluated a model using only the source database feature (Fig 3), and observed it to be significantly inferior, yielding a ROCAUC score of just 0.55, indicating that source is also not sufficient to explain the model’s performance. The same findings held for our other evaluated baselines.

Novel disease subtypes partitioning represents a challenging problem for both clinical and scientometric researchers. Despite the high quality medical ontologies already integrated into OT database, many diseases with similar symptoms may result from different causes, such as with pleiotropic genetic diseases, but they may not be annotated as such, even when pleiotropy is known (but their subtypes are not well defined), especially when the subtypes of pleiotropy are ambiguous [45]–[47]. This creates challenges when using OT to retrieve missing target-indication hypotheses due to the absence of direct candidates. While genetic association evidence on target-disease pairs can offer insights into relatedness [24], for our targets we lack actual negatives, or even a proxy measure such as annotation quality.

When awareness of possible subtypes exists, their identification and validation is currently manual, demanding exhaustive work by experts, who must also propagate their work into existing knowledge bases while drumming up awareness and consensus. Diseases common in developed countries, where clinicians have the resources to work with researchers may be more likely to be distinct, as opposed to neglected diseases in economically disadvantaged countries where doctors may not have the capacity to get their work published [48]–[50]. This issue may be worse for rare orphan diseases, which may have only a handful of dedicated researchers, reducing the chances of distinct manifestations being recognized and correctly annotated in knowledge bases.

To date, these limitations have restricted disease subtype discovery to a purely manual process, motivating our novel approach. We integrate OT direct evidence about each disease, including genetic, physiological and clinical features which are evaluated for prediction of which diseases have subtypes using machine learning models. We integrate unique features for each disease to represent their underlying properties, enhancing the identification of novel pleiotropies or overlooked annotations. Ultimately, our model produces a ranked shortlist of both new and potentially misclassified subtypes, which can then be validated by domain experts (S5).

### 3.1 Potential Implications

The methodologies in our study, including the combination of machine learning with OT, could extend to broader works. In clinical diagnostics; the increased identification and annotation of disease subtypes could provide better diagnoses, disease progression tracking and more personalized treatments and enhanced patient outcomes. In drug development, a nuanced understanding of disease subtypes can significantly benefit pharmaceutical research. By pinpointing specific diseases likely to have diverse pathologies, therapies and druggable, can be targeted with greater precision.

### 3.2 Conclusions

Annotating disease subtypes is crucial for enhancing our understanding of pathology and refining therapeutic strategies. By delineating diseases into subtypes, we pave the way for targeted research and treatment. We show that known disease subtypes can be mostly characterized automatically, that several diseases are likely to have uncharacterized subtypes, and a stability approach to identify them as a prelude to expert refinement and confirmation.

## 4. Methods

### 4.1. Overview Processing of Open Targets Dataset

Data was downloaded from Open Targets, as of July 2022 (7.22). The primary OT data sets used were associationByOverallDirect, diseaseToPhenotype, associationByDatasourceDirect, diseases. The subtype target was defined using the OT diseases dataset, according to whether a disease has any child links (“has_children”). The overall distribution of subtypes (“children”) across diseases is shown in **Fig 7**. We kept only direct associations, as we determined that indirect associations may leak target information.

**Fig 7.**
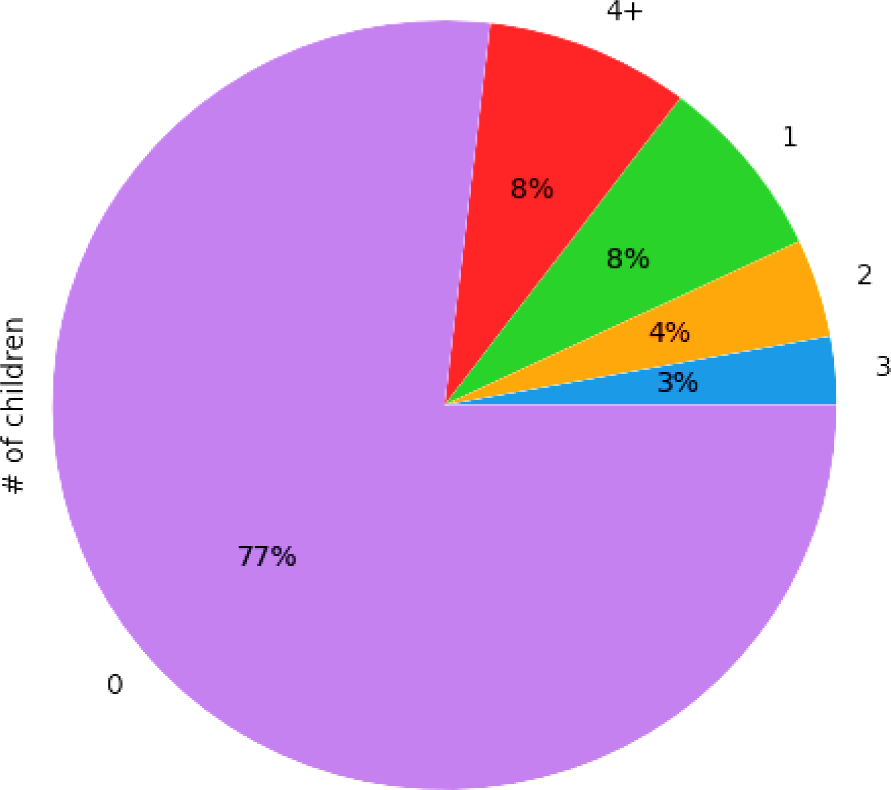
-. Subtypes per disease in Open Targets. The number of ‘children’ is shown. For 77% of the diseases, no partition to subtypes were recorded (marked as 0).

The initial dataset held 23,074 candidate diseases. We removed 2,817 irrelevant “diseases” relating to lab measurements (e.g., “IgG index” “BMI”). Diseases with the same name were aggregated together, with any “positive” target label taking precedence. 6,643 (32.8%) of disease terms have a known subtype. 3,035 terms with near-identical names (after removing white-space and lower casing) were merged, with positive subtype label taking precedence. The final modelling dataset held 17,222 diseases, of which 5,848 (34%) have at least 1 known subtypes. The temporal validation dataset uses a snapshot of the OT diseases table from 12.2023

### 4.2. Model Training and Evaluation

For most models, default hyperparameters were employed, with mean imputation of missing variables. For the tree models, we adjusted training class weights loss using the “square root balanced” hyperparameter. Logistic regression, K-nearest neighbours (KNN), linear support vector machine, histogram gradient boosting and random forest models were implemented using scikit-learn [31]. CatBoost, a boosting tree model, used the library of the same name. Features with a variance lower than 5e-4 were dropped. Tree models were used and favoured due to their speed, interpretability and historically superior performance on tabular data tasks. We also found that the tree models had the best performance on the task, as expected [19].

To evaluate the prediction of known disease subtypes, we employed stratified 5-fold cross-validation across all data points. In each iteration, the dataset was divided into training and testing sets, comprising 80% and 20% of the data, respectively. After training the model on the training set, predictions were made on the test set and recorded. The overall results were then assessed based on the accumulated test split outcomes. We also explored stratifying splits by ancestor disease groups, ensuring that training and testing sets did not share diseases with a common high-level ancestor. This was done to minimize bias from known diseases. Interestingly, this stratification had a marginal impact on performance, with the disease-stratified setup registering a 1% *increase* in absolute terms. Given these unexpected findings, we opted for the baseline split over the disease-stratified approach in all configurations, especially considering our consolidation by disease name.

Shapley (SHapley Additive exPlanations) values are used for summarizing feature importance to the trained model [51]. SHAP values are a popular method for interpreting feature importance, both globally, for specific data partitions or explaining individual predictions. It is used to show the relative contribution of each feature to the model’s output, also taking into account the contribution of other features to the model.

### 4.3. Generation of New Features

Features were extracted from OT direct evidence data sources. These included indicators of disease associated phenotypes and genomic transcript targets, and evidence scores per association. For example, the feature “Genetic association” refers to the evidence score from genetic association sources, “Literature” to literature evidence, “animal models” to the amount of evidence for a disease based on animal studies, and so on. For computational reasons, the genetic associations and phenotype sources were filtered to keep only those appearing at least 30 times in the dataset, and these were used as features, including their evidence scores. A novel approach [19], [36] of compressing these sparse features using a truncated singular matrix decomposition representation of ∼512 dimensions worked well in terms of performance and compute (not shown), but reduced interpretability, and thus was not used in the final model analysis. “Phenotype counts” is the number of distinct phenotypes associated with a disease (regardless of individual phenotype frequency or evidence score). “Max Phenotype frequency” is the overall frequency of the associated phenotype with the highest frequency in the data.

Engineered and aggregated features were extracted from the sources, including aggregated statistics (e.g., value mean, max, min, standard deviation, number of unique values, count of total occurrences)[19], [52]. The relative ratios of each evidence source type in relation to others was also extracted, e.g., the fraction of total evidence for a disease based on each type of evidence-source, and if a specific source was the largest or smallest ranked source (e.g., the feature “Literature ratio to biggest” is the amount of evidence from the Literature divided by the largest evidence source for the disease, which can also be the literature). For each disease we extracted the number of evidence counts per disease, per data source and data type, as well as additional features from the “disease” data including the total number of therapeutic areas, the existence of synonyms for a disease term, the number of direct parents, siblings (sharing the same direct parent) and ancestors for a disease in the OT graph, as well as the difference and ratio between the 2 features: “Ancestors sub parents” - the difference between the total number of ancestors and the number of direct parents for a disease. “Average associated evidence score” is the average confidence score of all evidences associated with a disease from all sources.

Overall disease population prevalence is estimated using the UK Biobank (UKB) [40]. The UKB contains demographic, lifestyle and medical information for 500,000 UK citizens. We matched 8,445 diseases to 663 ICD-10 medical diagnosis codes in the UKB, using Data-Field 41202 - “ICD10 diagnoses”. We crossmatched this with the overall frequency of these codes in the population as a feature used by the models. This feature did not contribute to model performance, and mainly served to help disprove whether diseases’ overall population prevalence rate might be a strong, potentially confounding feature [53].

### 4.4. Deep Learning Text Features

Using the state of the art techniques introduced in recent works combining tabular and pre-trained language models [19], [35], [54], [55], we used deep learning large language model pretrained on biomedical concepts, BioLORD-STAMB2-v1[56], to derive embedding features for each disease using its name and description. In brief, a pretrained neural network language model, trained to predict masked words in a text is taken, and the outputs from its final output layer is extracted and averaged across each token in the text. This mean-pooled output is used for features. Thus, texts are embedded in a vector space such that semantically similar text is close. We tried additional sentence-transformer language models, including all-MiniLM-L12-v2 [64], BioLord-2023 [65], BGE-en-base, and GTE-en-base [67], but their performance was slightly inferior (87∼88 AUC, not shown).

This approach lets us combine the benefits of large language models and deep representations in a simple, scalable way with our own features, while reducing possible name bias (e.g., diseases called “syndrome 1” - which could result in overfitting from a token-level finetuned language model) [55], [57]. These features are denoted as “Text Embedding X’’ in Fig 3, where X represents a vector in the embedding. For interpretability, we implemented an automated explanation framework showing exemplars of high, neutral and low values per embedding dimensions (Table 3), inspired by approaches in automated-machine learning works[35], [36], [52]. It is available in our codebase.

## Supporting information

Evaluation metrics and multiple model results on known subtypes prediction

- Novel candidate predictions, including ground truth and novel predictions and features for the 1531 cases where predictions differ from ground truth

- Literature search results for 800 highest confidence model predictions (known candidates model)

- Literature search results for top predicted novel candidate

## 5. Data Availability

Datasets used in the study are available on Open Targets: https://www.opentargets.org.

Code and results available online: https://github.com/ddofer/OpenTargets-DiseaseSubtype

## List of Abbreviations

CV: Cross validation. EFO: Experimental factor ontology. GxE: Gene x environment. GO: Gene ontology. GWAS: Genome-wide association studies. HPO: Human phenotype ontology. LR: Logistic regression.

OBA: Ontology of biological attributes. OT: Open Targets. PRAUC: Area under the precision-recall curve. ROCAUC: Receiver operating characteristic area under the curve. RF: Random Forest. UKB: United Kingdom BioBank. SHAP: Shapley additive explanations.

## Data Availability

All data available at https://github.com/ddofer/OpenTargets-DiseaseSubtype

